# The role of NIH funding in vaccine readiness; foundational research and NIH funding underlying candidate SARS-CoV-2 vaccines

**DOI:** 10.1101/2020.09.08.20187559

**Authors:** Anthony E Kiszewski, Ekaterina Galkina Cleary, Matthew J Jackson, Fred D Ledley

**Author notes:** Address reprint requests to Dr. Ledley at the Center for Integration of Science and Industry, Bentley University, 175 Forest Street, Waltham, MA, USA 02452 or. Author contributions: A.E.K, E.G.C., M.J.J., and F.D.L. designed research; A.E.K, E.G.C., M.J.J., and F.D.L performed research; A.E.K., E.G.C., M.J.J., and F.D.L analyzed data; and A.E.K, E.G.C., M.J.J., and F.D.L. wrote the paper; A.E.K. and F.D.L. supervised the project; F.D.L. acquired financial support.

## Abstract

This work characterizes the NIH contribution to vaccine technologies being employed in “warp speed” development of vaccines for COVID-19, as well as the lack of sustained NIH funding for published research against recognized epidemic threats. Using quantitative methods, we examined the advance of published research on ten of the vaccine technologies incorporated in the 165 candidate vaccines entering development through July 2020 as well as the NIH funding that supported this research. Live, attenuated virus, inactivated virus, and adjuvant technologies have been used in successful products since the 1950s and continue to exhibit steady advance. Synthetic (recombinant) vaccines, viral vectors, DNA, and TLR9 agonists as adjuvants emerged since the 1980s, and exhibit a logistic, “S-curve” pattern of growth characteristic of emerging technologies that have passed an analytically-defined *established* point. In contrast, mRNA, virus-like particle, and nanoparticle technologies show exponential growth characteristic of technologies short of their *established* points. The body of research and NIH funding for established and emerging vaccine technologies exhibited sustained growth through the late 2010s, supported by > 16,000 project years of NIH funding totaling over $17.2 billion (2000–2019), the majority through cooperative agreements and intramural programs. NIH funding for published research on vaccines for recognized zoonotic threats including coronavirus, Zika, Ebola, and dengue, however, has been inconsistent and reactive to disease outbreaks. These data are considered in the context of the high failure rate for candidate vaccines and evidence that technological maturity is a significant factor in the efficiency of product development. Sustained funding for both enabling technologies and vaccine development is essential to ensure a rapid response to COVID and future pandemic threats.

**SIGNIFICANCE STATEMENT:** This work examines the advance of research and NIH funding for technologies being employed in “warp speed” development of COVID-19 vaccines in the context of evidence that mature technologies have a greater likelihood of generating successful products. We show that candidate vaccines for COVID-19 employ a variety of established and still-emerging technologies, and identify $17.2 billion in NIH funding for this research from 2000–2019. In contrast, NIH funding for published research for vaccines on recognized pandemic threats has been inconsistent. This work highlights the significance and scale of the NIH contribution to vaccine technologies and the lack of sustained initiatives for vaccine development.

## INTRODUCTION

The COVID-19 pandemic has triggered an unprecedented mobilization of global vaccine development efforts. With fatality rates approaching or exceeding 1% (Basu, 2020), the prospect of long-term sequelae in those who recover (Demertzis et al., 2020; Fiani et al., 2020), and the high level of population immunity required to halt transmission (Flaxman et al., 2020), initiatives are expected to proceed at “warp speed” (O’Callaghan, Blatz, & Offit, 2020; Corey et al., 2020).

Within six months of the first description of the SARS-CoV-2 virus (Zhu et al., 2020), a wide variety of vaccine candidates entered clinical development (WHO, 2020). Many were based on platforms such as attenuated or inactivated virus, synthetic (recombinant) proteins, and conventional adjuvants that have already been used in successful vaccines for diseases such as influenza or polio. Some exploited technologies previously shown to generate immune responses against other human coronaviruses, including Middle East Respiratory Syndrome virus (MERSCoV) (Modjarrad, 2016) and SARS-CoV-1 (McPherson et al., 2016). Some incorporated technologies utilized in vaccines registered for veterinary use against animal coronaviruses such as Infectious Bronchitis Virus (IBV) (Li et al., 2018) and bovine Betacoronavirus-1 (Decaro et al., 2009). Others incorporated novel technologies, such as recombinant viral vectors, DNA, or mRNA, which have not yet demonstrated safety and efficacy in humans.

Vaccine development remains daunting on several fronts despite a vaunted history that includes the global eradication of smallpox, the near elimination of polio (Plotkin, 2014), the emergence of effective vaccines for seasonal influenza, common childhood diseases, and many other viral and bacterial pathogens. Evidence suggests a failure rate of 94% for vaccines entering clinical development, and that, for successful products, the timeline from preclinical studies to approval is 10.7 years (Pronker et al., 2013). There are a number of reasons for this high failure rate and prolonged developmental timeline.

First, many pathogens have been refractory to vaccine development. High mutation rates render HIV a moving target (Safrit et al., 2016), while influenza undergoes rapid evolution through both antigenic shift and drift (Kim, Webster, & Webby, 2018). Strain differences trigger adverse immune responses with dengue virus (WHO, 2018), and extreme diversity and immunoevasive adaptions have complicated development of vaccines for malaria parasites (Laurens, 2018). Prior sensitization to non-TB bacterial antigens reduce the BCG vaccine’s impact on tuberculosis (Arregui et al., 2016).

Second, some vaccine development challenges are platform-based and independent of the target. Adverse activation of and interference by Type I interferon has plagued the development of RNA-based vaccines (Pardi et al., 2018; De Beuckelaer et al., 2016). DNA-based vaccines are prone to insufficient stimulation of immunity (Porter & Raviprakash, 2017).

Third, development of public health vaccines is classically challenged by issues of cost, stability, and safety. The market for public health vaccines is extremely cost-sensitive given limited public health budgets, the reliance of many regions on non-governmental organizations such as UNICEF or PAHO to purchase vaccines, and the need to vaccinate large numbers of people (Randolph & Barreiro, 2020) to achieve community protection. Moreover, many vaccine technologies produce thermally unstable products requiring refrigeration in lieu of rapid administration. This can dramatically increase distribution costs or render widespread use impractical (Kristensen et al., 2016).

Fourth, given the intense scrutiny of vaccine safety in light of misleading claims and public skepticism of vaccines fomented by the anti-vax movement, widespread public acceptance will require developers to demonstrate an exemplary margin of efficacy and safety (Ball, 2020). Finally, it should be noted that general public health measures can also be effective against pandemic disease, and the rapid dissipation of outbreaks related to viruses such as SARS-CoV-1 and Zika have made thorough clinical validation of vaccines impossible (Thomas & Barrett, 2020).

This work is predicated on innovation research across many different fields demonstrating how the maturation level or readiness of a technology is a critical determinant in its ability to generate products that successfully meet market needs (Christensen, 1992; Christensen, 1997; GAO 1999; Clausing & Holmes, 2010; Foster, 1982; McNamee & Ledley, 2012). Our previous work has explored the applicability of this principle to drug development using a bibliometric model for the maturation of basic, biomedical research on novel drug targets or therapeutic modalities based on the rate of publication (McNamee, Walsh, & Ledley, 2017). Studies on the development of more than 500 approved drugs have shown that few targeted therapeutics are successfully developed before research on both the drug target and the therapeutic modality have passed an analytically derived *established* point, and that the timelines of development are significantly shorter when clinical trials commence after this point (Beierlein et al., 2017, McNamee & Ledley, 2017, McNamee, Walsh, & Ledley, 2017, Ledley et al., 2014, Cleary, Jackson, & Ledley, 2020).

Additional studies have examined the scale of NIH funding for this body of enabling, basic research (Cleary et al., 2018; Cleary, Jackson, & Ledley, 2020). These studies show that the NIH contributed more than $230 billion in funding for research related to the 356 drugs approved from 2010–2019 or their 218 drug targets. Significantly, more than 85% of this funding was categorized as basic research on the drug targets, rather than research on the drugs themselves.

The present work examines the maturation of the technologies employed in current efforts to develop COVID-19 vaccines. Specifically, we examine the timelines of published research on technologies related to adjuvants, recombinant proteins, nucleic acids, attenuated/inactivated virus, viral vectors, and formulations using virus-like particles or nanoparticles, the maturity of this research at the end of 2019, and the NIH funding that contributed to this research. We also examine the timelines of research directed towards vaccines for coronaviruses and three unrelated viral pathogens that have been associated with epidemic transmission (Zika, Ebola, and dengue). We consider the impact of this prior research on accelerated efforts to develop a vaccine for COVID-19, and the importance of sustained public sector funding in establishing a foundation for responding to pandemic outbreaks.

## RESULTS

### Research on vaccine technologies

As of July 31, 2020, the WHO listed 165 candidate vaccines against COVID-19: six in Phase 3 trials, twenty in Phase 1 and/or 2, and 139 in various stages of preclinical development (WHO, 2020). PubMed searches were performed for ten prevalent vaccine technologies from the portfolio of products described in the WHO report. The technologies examined and number of research publications related to each topic are shown in Table 1. Cumulative publications over time are shown in Figure 1A.

**Table 1.**
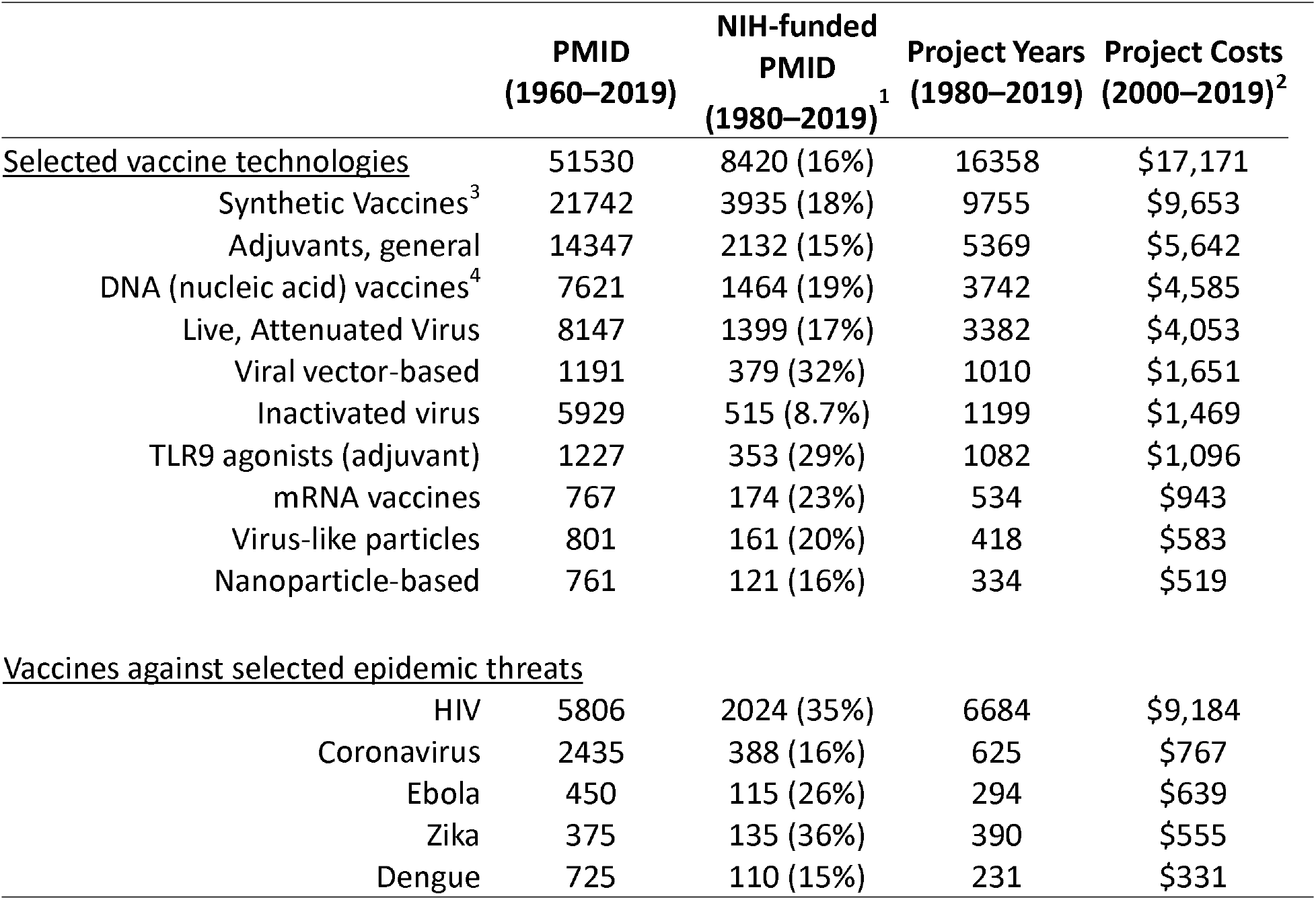
Publications in PubMed (PMID) and NIH funding associated with research on technologies used in candidate COVID-19 vaccines as well as vaccine development for selected epidemic threats.

**Figure 1.**
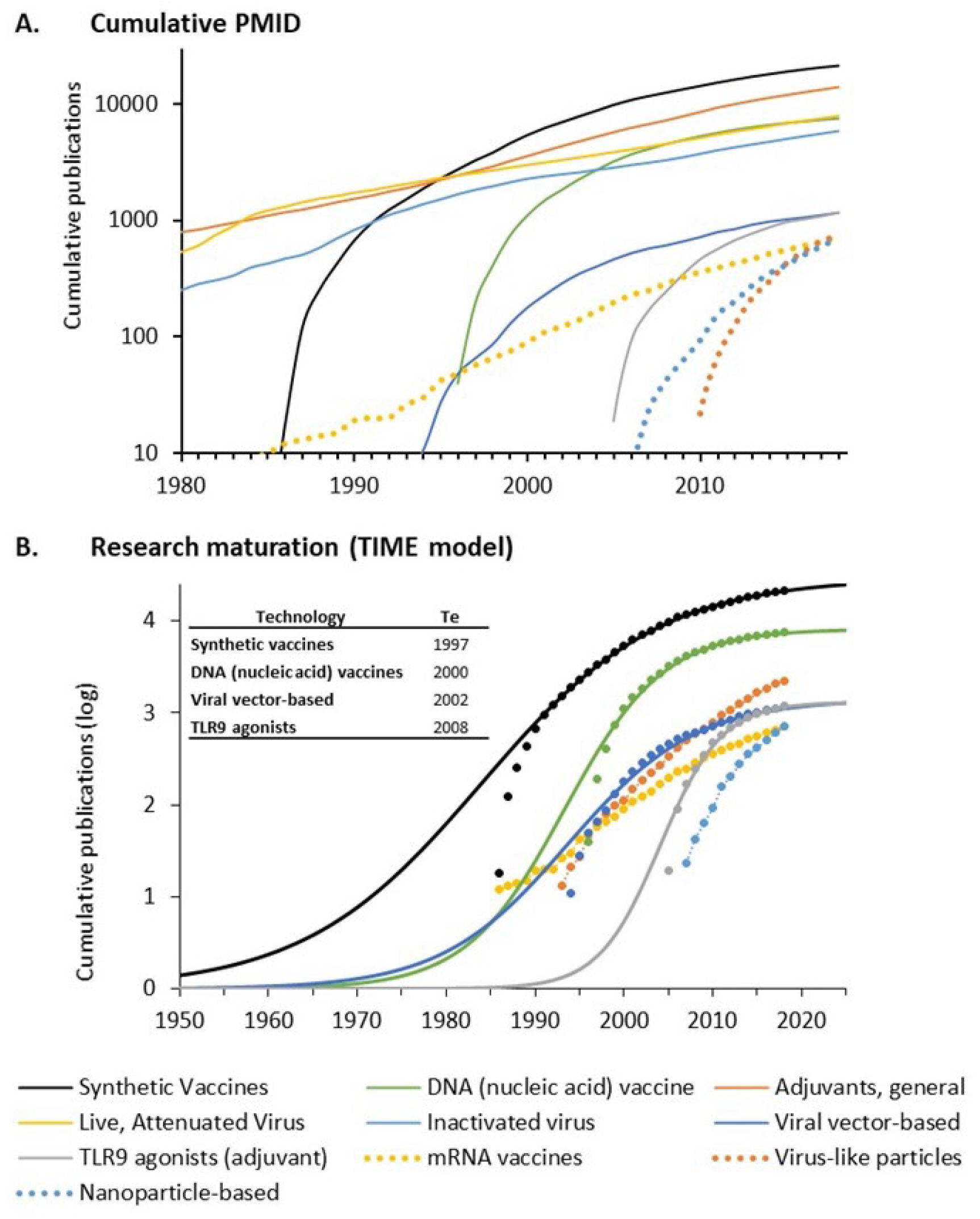
Publications related to ten vaccine technologies used in candidate COVID-19 vaccines. A. Cumulative PMID from PubMed search over time. Data is shown on a logarithmic scale. B. Maturation of research using the TIME model. Cumulative publications are shown as symbols. Curves fitting the (exponentiated logistic) TIME model are shown as solid lines. The calculated established points for thesetechnologies are shown in the box. Curves exhibiting exponential advance for technologies that have not yet reached an established point are shown as dotted lines. Supplemental Figures 1–3 provide additional curve fits.

Several viral technologies involving whole virus preparations, including live/attenuated and killed/inactivated viral vaccines, as well as non-specific adjuvants, predate the modern era. These are used in classical vaccines including polio, influenzas, MMR, and others that emerged as prominent elements of public health in the 1950s. Research on these technologies has accumulated steadily since 1980 (Figure 1A). Inactivated viral approaches accounted for four of the first ten candidate vaccines to enter human clinical trials against COVID-19, but only 12(7.3%) candidate vaccines as of July 31, 2020.

Synthetic vaccines employing recombinant protein subunits emerged in the 1980s, capitalizing on the advances in large-scale protein production and purification. Exponential growth of research on synthetic vaccines was evident after 1985 followed by steady growth from the mid-1990s to the present time (Figure 1A). While only one of the first ten COVID-19 candidate vaccines to enter clinical trials employed these technologies, recombinant protein antigens are incorporated in 66 (40%) of candidate vaccines as of July 31, 2020.

Vaccines based on recombinant viral vectors that express immunogens emerged in the mid-1990s as an outgrowth of research on gene therapy and experienced a period of exponential growth and subsequent slowing from the early 2000s to the present (Figure 1A). Two of the first ten COVID-19 candidate vaccines to enter human trials were based on recombinant adenoviral vectors. Other vaccines currently in clinical development use other viral vectors, including VSV, measles, Sendai virus, avian paramyxovirus, baculovirus, lentivirus, and influenza. Overall, viral vectors are employed in 31 (19%) candidate vaccines.

DNA-based vaccine technologies emerged in the mid-1990s, also as an outgrowth of research on gene therapy, and exhibited a period of rapid acceleration and subsequent slowing. These technologies involve injection of plasmid or synthetic DNA molecules, with the expectation that the injected DNA will be taken up by cells in the body and express the antigen.

A related nucleic acid-based approach involves direct injection of mRNA, which cells take up and translate into antigenic proteins. This approach derives from early methods for expressing in vitro translated mRNA in *Xenopus* oocytes, and production of infectious virus through injection of purified viral mRNA into cultured cells (Schlake et al., 2012). This field has exhibited steady growth with the emergence of advanced nucleotide chemistries, self-replicating mRNA, and improved delivery methods.

One DNA vaccine and two mRNA vaccines were among the earliest candidate vaccines to enter human trials. Both mRNA vaccines were among the first to enter Phase 3 clinical evaluation. Overall, there were 36 (22%) nucleic acid-based vaccines in development as of July 31, 2020, 14 based on DNA and 22 with RNA.

Several other vaccine technologies currently employed in candidate COVID-19 vaccines are also shown in Figure 1A, including lipid nanoparticles and virus-like particles designed to enhance delivery and antigenicity of recombinant proteins as well as the use of novel adjuvants (i.e., toll-like receptor 9 [TLR9] agonists) designed to stimulate specific immune pathways. These technologies were employed in 24 (15%) of the 165 candidate vaccines under development on July 31, 2020.

The maturity of the seven technologies emerging since 1980 was examined using the Technology Innovation Maturation Evaluation (TIME) model (Figure 1B). This model fits the cumulative number of publications to an exponentiated logistic function. Plotted on a log scale, this function exhibits the logistic “S-curve” that is characteristic of technology growth in many different fields, including drug development (Foster, 1982; Christensen, 1992; Christensen, 1997; McNamee & Ledley, 2012; McNamee, Walsh, & Ledley, 2017). From this equation, it is possible to calculate the *initiation* point of research on each topic (defined as the point of maximum acceleration) and the *established* (Te) point (defined as the point of maximum deceleration) (Figure 1B). Individual curve fits are shown in Supplemental Figures 1, 2, and 3.

Four technologies (synthetic, DNA, viral vector, and TLR9 agonists) exhibited a logistic pattern of growth characteristic of maturity (Figure 1B). Each of these technologies passed their *established* point prior to 2010. Three technologies (virus-like particles, mRNA, and nanoparticles) did not exhibit a logistic pattern of growth with clear evidence of maximal deceleration. This pattern is characteristic of technologies that have not yet passed their *established* point.

### NIH funding for published research on vaccine technologies

The NIH RePORTER database was used to identify published research (via PMID) acknowledging NIH funding support, specific projects and project years associated with these publications, and the project costs for those years. The total numbers of PMID, number and percent NIH-funded PMID, project years, and project costs for ten vaccine technologies are shown in Table 1. We identified a total of 51,530 publications related to these ten vaccine technologies, including 8,420 (16%) associated with NIH funding, comprising 16,358 project years and $17.2 billion in project costs. The NIH contributed the most funding for synthetic (recombinant) vaccines ($9.65 billion) followed by adjuvant research ($5.6 billion), DNA vaccines ($4.6 billion), and live, attenuated virus ($4 billion). More than $1 billion in NIH funding was associated with research on inactivated virus and viral vector-based vaccines as well as TLR9 agonists as adjuvants. More than $500 million was invested in mRNA vaccines, virus-like particles, and nanoparticle-based vaccines combined.

The annual number of publications, NIH-funded publications, project years, and project costs for the ten technologies are shown in Figures 2A–B. The annual totals for each technology individually are shown in the interactive graphic https://tabsoft.co/31EkYeK. Publication activity increased steadily from 1980 to 2010, with the fraction of publications acknowledging NIH funding rising from 4% in 1980 to 20% in 2010. After 2010, annual publications and the fraction of NIH-funded publications both decreased. The number of NIH project years and costs similarly increased from 2000–2010, and have decreased considerably in recent years.

**Figure 2.**
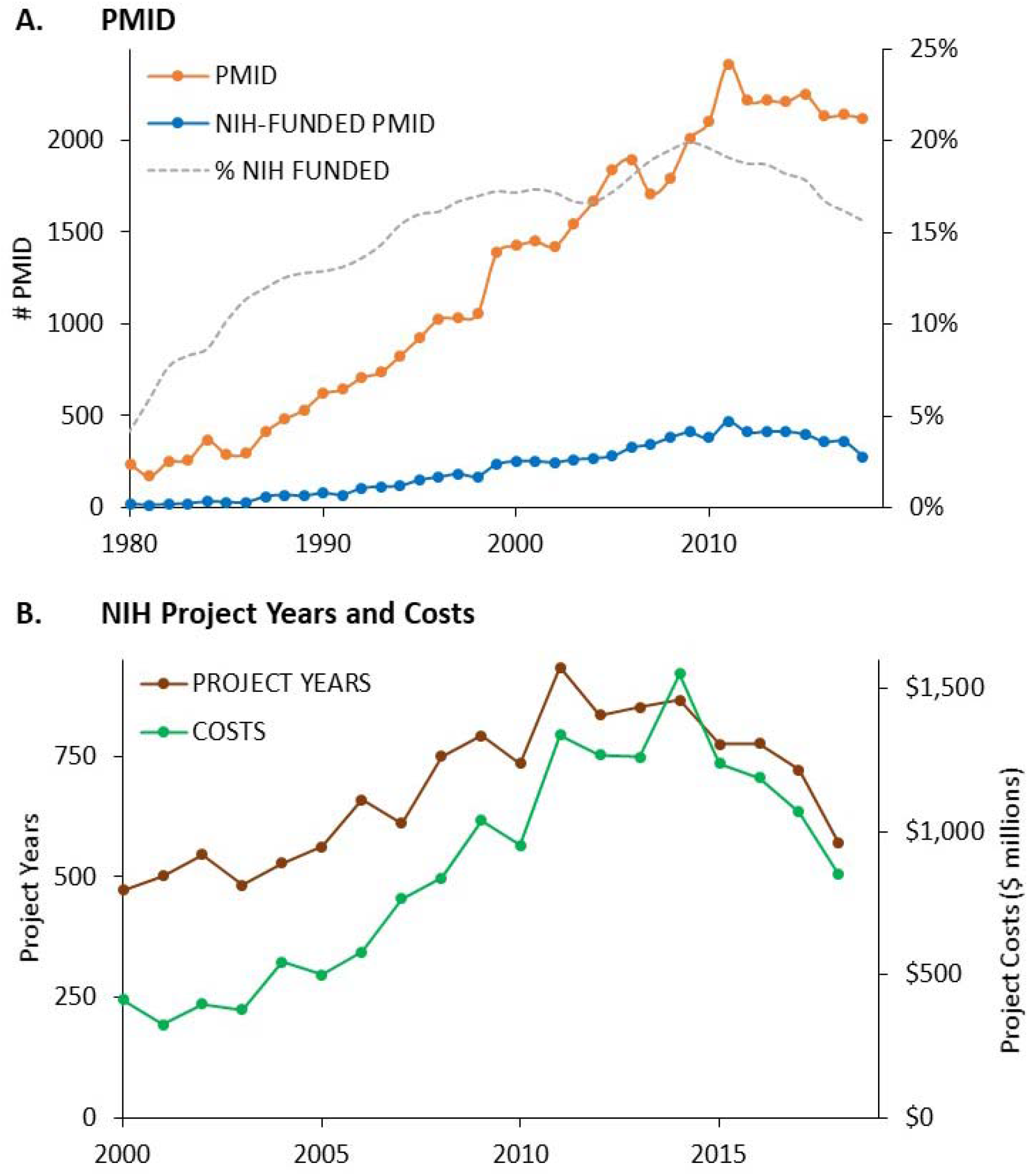
NIH support for published research on ten vaccine technologies used in candidate COVID-19 vaccines. A. Annual PMID, NIH-funded PMID, and the fraction of PMID receiving NIH support for vaccine technologies. B. Annual project years and project costs associated with NIH-funded PMID 2000–2019.

The activity code categories for NIH projects associated with published research on vaccine technologies are shown in Figure 3A. While 40% of project years associated with research on these technologies represented investigator-initiated research projects, these accounted for only 8.9% of the total project costs. A greater fraction of funding was associated with cooperative agreements (44%), intramural programs (9.4%), and research program projects and centers(22.7%). This pattern differs sharply from that seen for development of new drugs approved from 2010–2019 (Cleary, Jackson, & Ledley, 2020), where the majority of project years and funding for research on drug targets was associated with investigator-initiated research projects. However, this pattern does resemble that of molecular research underlying the discovery and development of remdesivir for treatment of COVID-19 (Cleary, Jackson, Folchman-Wagner & Ledley, 2020).

**Figure 3.**
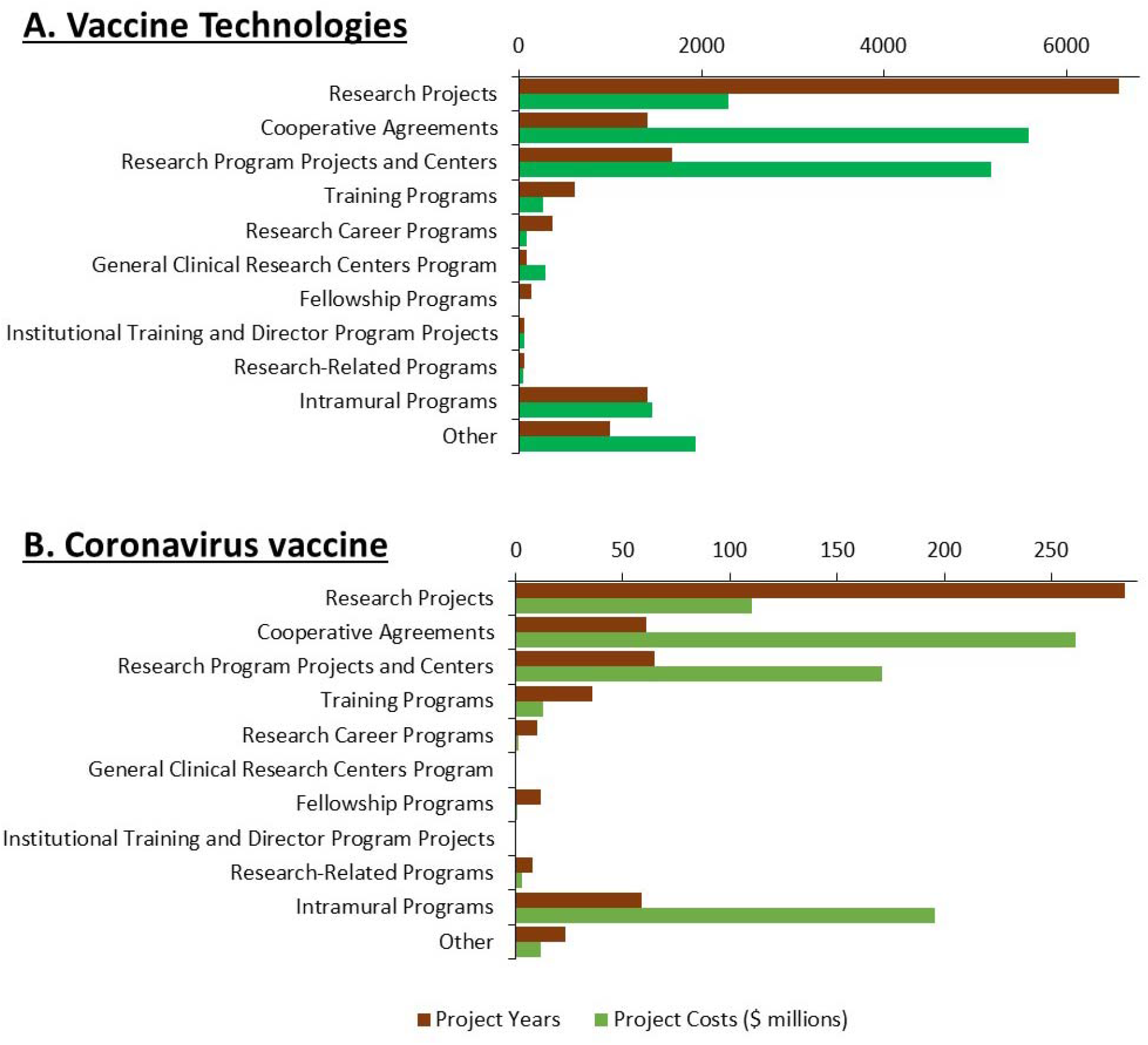
NIH funding for published research on ten vaccine technologies and coronavirus vaccines by activity category. A. Number of project years and project costs for research on ten vaccine technologies by activity category 2000–2019. B. Number of project years and project costs for research on coronavirus vaccines 2000–2019.

### Research and NIH funding for diseases with pandemic potential

Table 1 shows the number of PMID, NIH-funded PMID, project years, and project costs associated with vaccine development for HIV, which has been the subject of intensive vaccine research since the 1980s, as well as Ebola, Zika, dengue, and coronavirus. Figure 4A shows the timeline of combined NIH project costs for these four viruses since 2000 compared to the project costs for research on HIV vaccines. NIH funding for research on HIV vaccines increased more than fourfold from 2000–2014, and has subsequently declined. There was little NIH funding for research on Ebola, Zika, dengue, or coronaviruses prior to 2014.

**Figure 4.**
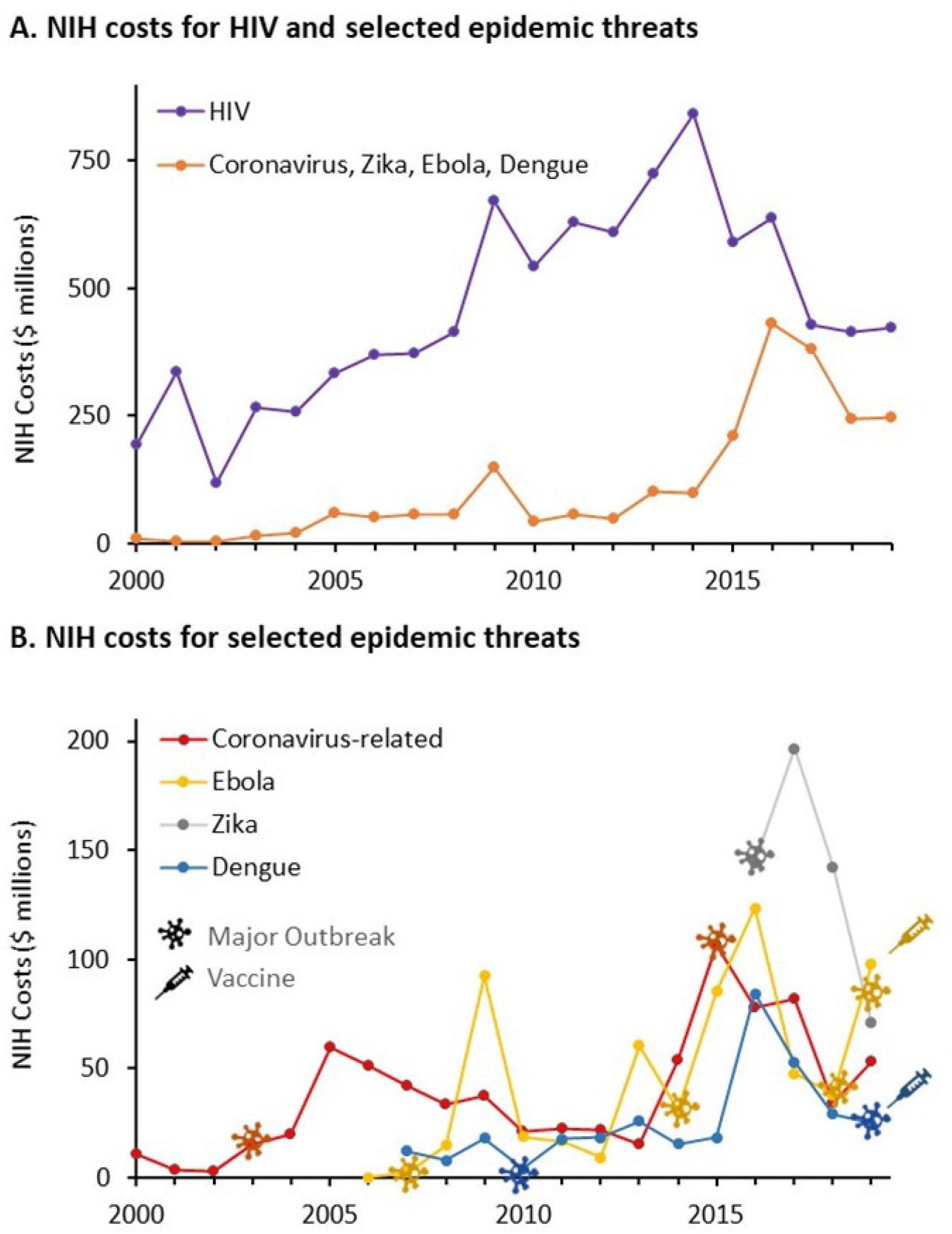
NIH costs for published search on vaccines for HIV and selected zoonotic pandemic threats (Coronavirus, Ebola, Zika, dengue) 2000–2019. A. Annual NIH costs associated with published research on HIV vaccines compared to the cumulative total of research on Coronavirus, Zika, Ebola, and dengue. B. Annual costs associated with published research on Coronavirus, Zika, Ebola, and dengue. Symbols indicate years of major outbreaks as well as the years of vaccine approval.

Figure 4B shows the annual project costs for vaccine research on Ebola, Zika, dengue, and coronaviruses individually. The annual totals for PMID, NIH-funded PMID, projectyears and projects costs for research related to each virus is shown in the interactive graphic https://tabsoft.co/31EkYeK. NIH funding associated with publications on vaccines for Ebola virus exhibited a sharp peak in 2009, a larger peak from 2013–15, and another resurgence in 2019. These peaks corresponded to a relatively small outbreak of Ebola in 2007, the major outbreak in 2014, which had limited spread to Europe and the US, and an ongoing event that began in 2018 (https://www.cdc.gov/vhf/ebola/history/chronology.html).

A single peak of NIH funding associated with publications related to vaccines for Zika was evident, corresponding to the 2015–17 outbreak in South America and the Caribbean that also impacted several US states. Another funding peak was evident following the outbreak of SARSCoV-1 in Asia and its subsequent spread to Toronto. A rise in funding for publications related to dengue was apparent in 2016, coincident with recognition of the increasing prevalence of this disease, but dropped rapidly coincident with an abrupt decrease in reported cases in 2017 and 2018 (https://www.who.int/news-room/fact-sheets/detail/dengue-and-severe-dengue). These data indicated a lack of sustained NIH funding for vaccine research in the wake of waning disease outbreaks.

The pattern of NIH funding for published research on coronavirus vaccines shows a similar lack of sustained funding and research activity. NIH project costs associated with published research on coronaviruses peaked in 2005, then dropped steadily to 2013. A second peak of NIH funding associated with published research on coronavirus vaccines was evident following the MERS outbreak in 2012, which peaked in 2015 and then declined rapidly.

Figure 3B shows the activity code categories for the NIH projects associated with published research on coronavirus vaccines. While the majority of project years were associated with investigator-initiated research projects, only 14% of project costs were associated with research projects, while 34% went to cooperative agreements, 26% to intramural programs, and 22% to research program projects and centers. Thus, as with NIH funding for research on vaccine technologies, the majority of project costs came through research organized or conducted by government agencies, rather than from investigator-initiated projects.

## DISCUSSION

This analysis was undertaken in the context of research showing that there is a 94% failure rate for vaccines entering development (Pronker et al., 2013). The exigency for “warp speed” development of a COVID-19 vaccine to stem the ongoing pandemic (O’Callaghan, Blatz, & Offit, 2020; Corey et al., 2020), and evidence that maturity of the underlying technologies is a significant factor in the efficiency of biopharmaceutical development (McNamee, Walsh, & Ledley, 2017; Cleary, Jackson, & Ledley, 2020). Using quantitative methods, we examine published research on ten technologies that are being used in candidate vaccines for COVID-19 and estimate the maturity, or readiness, of these technologies for clinical development. We also describe the NIH contribution to development of these technologies through the end of 2019, as well as support for vaccine development directed towards specific zoonotic threats like coronavirus, Zika, Ebola, and dengue.

Some of the technologies used by COVID-19 candidate vaccines (live attenuated or inactivated virus, non-specific adjuvants) date from the mid-20^th^ century and have been used to create many successful vaccine products. While we found that the research literature on these technologies continues to grow, there is no evidence of exponential acceleration since 1980, suggesting that these technologies had become well-established before that time.

Research on synthetic (recombinant) vaccines, DNA vaccines, (recombinant) viral vectors, and TLR9 agonist-based adjuvants exhibited an “S-curve” pattern of publication activity when fitted to the TIME model. Their curves’ inflection points suggest that each of these technologies was *established* prior to 2010. In contrast, three technologies—mRNA vaccines, virus-like particles, and nanoparticles—exhibit exponential patterns of growth through 2019, suggesting these technologies have not yet reached their *established* point.

It should be emphasized that the TIME model does not attempt to identify singular publications representing significant milestones in vaccine development. Rather, the model posits that the complete body of published research describing original insights or inventions, the replication or refinement of previous observations, and/or refutation of erroneous results all contribute to the foundation of knowledge required for efficient product development.

Previous research using the TIME model has demonstrated a relationship between metrics of technology maturation and the efficiency of development. Studies of drug development for cancer (McNamee & Ledley, 2017) and cardiovascular disease (Beierlein et al., 2017) demonstrate that few new drugs were approved before research on the biological target or class of drugs passed the *established* point. Similarly, approvals were rare for products utilizing monoclonal antibodies (McNamee & Ledley, 2012), nucleotide therapeutics (Beierlein, McNamee, & Ledley, 2017), and gene therapies (Ledley et al., 2014) before research on these modalities passed the *established* point. Moreover, the timeline of clinical development from Phase 1 to approval is significantly shorter when clinical trials commence after the underlying technologies have passed this point (McNamee, Walsh, & Ledley, 2017;Cleary, Jackson, & Ledley, 2020; Beierlein et al., 2017). These observations are consistent with studies of technological innovation in other sectors, which show that nascent technologies are commonly unable to meet the standards required by established markets, despite the fact that these technologies offer potential advantages and may ultimately supplant incumbent technologies in the marketplace (Christensen, 1992; Christensen, 1997; McNamee & Ledley, 2012).

Experience with vaccine development offers many parallels. The success of the Salk vaccine was preceded by several unsuccessful efforts to develop inactivated or attenuated viral vaccines. Moreover, the roll-out of the Salk vaccine itself was complicated by an outbreak of polio caused by inadequate inactivation of early batches of the product, which led to temporary suspension of the vaccine program months after the vaccine was approved (Blume, 2005). Similarly, early versions of both RSV and Measles vaccines were associated with enhanced disease on reexposure, reflecting the immature knowledge of vaccines and the immune system in that era (Lambert et al., 2020). More recently, a dramatic decline in the success rate for vaccine development from 2003–2013 has been ascribed, in part, to the repeated failures of then-nascent DNA technologies and unresolved problems with immunogenicity (Stephens, 2014). In contrast, the remarkable safety of vaccines currently approved by the FDA (Tau, Yahav, & Shepshelovich, 2020), as well as the annual development of seasonal influenza vaccines for emerging strains of influenza virus, are consistent with the efficiency of *established* vaccine technologies.

This pattern is incorporated in an economic model of vaccine development prepared for the Coalition for Epidemic Preparedness Innovations (CEPI), in which vaccine technologies having “no licensure track-record” are assumed to be associated with a greater risk of vaccine failure and higher development costs than “well-established” technologies (Gouglas et al., 2018). The CEPI model estimates the failure rate of vaccines in development is greater than 90%, and the cost of developing a vaccine from preclinical through Phase 2a (entry of Phase 3) to be between $137 million and $1.1 billion. This wide range reflects, in part, differences in the cost of development for technologies without, or with, a track record of licensure, as well as variations in indirect cost (Gouglas et al., 2018).

In this context, the over-representation of mRNA-based vaccines among the leading COVID-19 candidate vaccines is of some concern given the relative immaturity of these technologies as measured by the TIME model. Nucleic acid-based vaccines were able to move more rapidly into clinical trials (WHO, 2020) due to inherent efficiencies in process development and scale up of production of these products. This analysis suggests that there may be substantially more downstream risk of product failure or delay with such immature technologies.

We would also note that there is no licensure track record for adenoviral vaccines, which were also able to rapidly enter clinical trials and progress through the initial development stages. Prior to the emergence of COVID-19, two MERS vaccines utilizing these technologies passed Phase 1 trials before the waning of these diseases precluded further testing (Modjarrad et al., 2019; Folegatti et al., 2020).

The cost of vaccine discovery and development typically relies on both public and private sector investments. A 1997 report by the National Vaccine Advisory Committee concluded that “two thirds of all new vaccines provided worldwide have been produced by a US network of independent industrial, governmental, and academic partners engaged in vaccine research and development” (NVAC, 1997). A 2001 report by the World Bank, working with Gavi, the Vaccine Alliance, described the role of public sector institutions as being primarily focused on basic science “measured by the number and value of the research manuscripts they publish in the scientific literature” (Batson & Bekier, 2001; Francis, 2010). In contrast, industry focused on development, manufacture, and marketing of vaccine products (Batson & Bekier, 2001), and more than 90% of all vaccine doses procured in the past five years have been manufactured by for-profit organizations (WHO, 2019).

We also examined NIH funding related to four diseases with recognized epidemic potential including coronavirus, Zika, Ebola, and dengue. The first three were identified by the WHO as pandemic risks in the 2016 “R&D Blueprint for Action to prevent Epidemics” (WHO, 2016) and have been identified as priority epidemic infectious diseases by CEPI (Gouglas et al., 2018). Dengue is an endemic pathogen that causes focal outbreaks, which was considered to be outside the scope of the 2016 Blueprint (Mehand et al., 2018).

A lack of sustainable investment has long been recognized as a limiting factor in developing vaccines for emergent pandemic threats, particularly those initially impacting low-income countries (Rappuoli, Black, & Bloom, 2019). This problem is exacerbated by the fact that vaccine development typically requires coordinated and sustained funding for research encompassing immunization technologies, disease pathogens, vaccine formulation, and clinical development.

Our analysis describes a sporadic pattern of grant funding related to coronavirus, Zika, Ebola, and dengue viruses, in which outbreaks of disease trigger an increase in grant-funded publication activity, which then wanes rapidly over several years. This is consistent with the observation that investigator-initiated grant mechanisms have not proved to be a robust mechanism for vaccine development (Rappuoli, Black, & Bloom, 2019).

Our work also demonstrates that while the largest fractions of PMIDs and project years represented investigator-initiated research projects, a majority of the funding was associated with government-initiated research through cooperative agreements or intramural research, or for research capacity through research program projects and centers. This pattern has been observed previously in examining the foundational research underlying development of remdesivir (Cleary, Jackson, Folchman-Wagner, & Ledley, 2020). This pattern, however, is distinctly different from that of NIH funding associated with drugs in other therapeutic areas, where the large majority of research funding involves investigator-initiated research projects or related research program projects and centers (Cleary, Jackson, & Ledley, 2020). Further research is required to understand the determinants of investigator-initiated versus government-initiated research, and how these different mechanisms contribute to the development of vaccine technologies and vaccines for pandemic threats.

Finally, we would note that there is no demonstrated relationship between the amount of NIH funding and the success of vaccine development initiatives. Despite decades of research and billions of dollars in NIH funding, vaccines have yet to be registered for diseases such as HIV and hepatitis C. In contrast, the NIH contributed little funding to published research on Ebola or dengue viruses, both of which are associated with FDA-approved vaccines.

## Limitations of this research

There are several important limitations to this study. First, this analysis is restricted to research that produces publications in PubMed, which may not capture research described in white papers, reports, patents, regulatory filings, unpublished clinical studies, or trade secrets. This method may also fail to identify publications describing enabling technologies not directly related to vaccines, early research performed before accepted vocabularies emerge, and publications without abstracts in PubMed. Second, it is not possible to associate costs with every NIH-funded publication due to missing data in RePORTER and inconsistencies between publication dates and project years. Moreover, given an estimated lag of up to three years between research funding and publication (Boyack & Jordan, 2011), funding for research performed since 2018 may be underrepresented in the data collection. Third, this analysis does not account for research funded by agencies outside HHS, particularly DOD or NSF, government contracts, other nations, philanthropies, or academic institutions. Nor does it account for the $1.8 billion that the U.S. contributed to Gavi since 2009 (KFF, 2020) or for investments by industry in clinical trials of process development.

## Conclusion

The “Operation Warp Speed” and “Accelerating COVID-19 Therapeutic Interventions and Vaccines (ACTIV)” initiatives involved funding a variety of different candidate vaccines utilizing different technologies. This work demonstrates that the technologies represented in this portfolio vary significantly in maturity. Inasmuch as technological maturity is often a significant factor in the efficiency and cost of product development, the maturity of individual technologies should be a factor in guiding both expectations for success and investment in different candidate vaccines.

This work highlights the scale and importance of a broad, sustained foundation of NIH funding in the ability to respond to the COVID-19 pandemic. The research that will enable rapid development of a COVID-19 vaccine was not undertaken with such a vaccine in mind, but was often undertaken in unrelated contexts such as basic molecular biology, gene therapy, immunology, or drug delivery. Nevertheless, this funding advanced the maturation of technologies that are now available for application in COVID-19 vaccines. Significantly, the majority of this funding came in the form of government-initiated cooperative agreements or intramural research, rather than investigator-initiated research projects.

In contrast, this work again demonstrates the lack of sustained research and NIH funding related to recognized, epidemic threats, such as coronavirus, may have contributed to the lack of preparedness for the emergence of COVID-19. Further consideration needs to be given to the mechanisms of funding research on emerging pandemic threats beyond coronavirus (WHO, 2016) to ensure that the technologies are ready to respond.

## METHODS

Technologies utilized in candidate vaccines against COVID-19 were identified from the WHO “DRAFT Landscape of Candidate COVID-19 vaccines” (WHO, 2020) accessed July 31, 2020. Viruses with epidemic potential that have been subject to intensive vaccine development initiatives were identified from Plotkin, 2017, and the WHO Blueprint (Plotkin 2017; Mehand et al., 2018).

Searches were performed in PubMed (https://www.pubmed.gov; accessed June 3, 2020) using the updated Automatic Term Mapping (May, 2020), initially with Medical Subject Headings (MeSH) terms, and then optimized with Boolean modifiers to increase specificity. PubMed Identifiers (PMIDs) and publication year were recorded for each publication identified.

We then quantified the contribution of NIH funding to publications identified in PubMed using NIH RePORTER (https://exporter.nih.gov/ExPORTER_Catalog.aspx; accessed June 3, 2020) via methods reported in a similar study of drug development (Cleary et al., 2018). A tabulation of NIH-sponsored research that led to these publications was derived from the RePORTER/ExPORTER format files catalog (NIH, 2020). Wherever possible, each PMID was associated with a project year corresponding to the project number and fiscal year in the “Link Tables for Project to Publication Associations.” The “Project Data Table” provided the fiscal year costs for each project (since 2000). Costs were derived for each project year corresponding to the program cost associated with PMIDs via the Project Table. Activity codes associated with the core project indicated the type of grant.

Publications occurring before the first year of the grant award or more than four years after the last year of the grant were excluded. Publications 1–4 years after the last year of the grant were associated with the project costs of the last year. Grant categories were derived from “NIH Types of Grant Programs 2020” (https://grants.nih.gov/grants/funding/funding_program.htm; accessed May, 2020). Costs are given in constant dollars inflation adjusted to 2018 using the U.S. Bureau of Labor Statistics All Urban Consumer Prices (Current Series) (https://www.bls.gov/data/; accessed May, 2020). All analyses were performed in PostgreSQL and Excel.

The relative maturity of seven critical technologies emerging since 1980 was examined by fitting the bibliographic Technology Innovation Maturation Evaluation (TIME) model, an exponentiated logistic function described previously, to a time series of publication records derived from PubMed searches (McNamee, Walsh, & Ledley, 2017). The equation has the form:

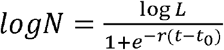

Where *N* is the number of publications, *L* is the presumed upper limit of publications, *r* is the growth rate, *t* is time, and *t*_0_ is midpoint of the exponential growth. This asymmetric sigmoidal function exhibits the common logistic sigmoid function over log scales. The parameters were fit to time-series publication data using a non-linear least squares implementation of the Levenberg-Marquardt algorithm (LMFIT, version 1.0.1).

The *initiation* and *established* points, representing the beginning and end of the exponential growth phase are defined as the points of maximum and minimum acceleration respectively, or log*N*’’(*t*)_max,min_. These points are analytically determined by:

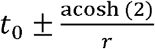

## Data Availability

All data are freely available on scholar@bentley (scholar.bentley.edu), part of the Digital Commons (bpress.com) platform.

## ACKNOWLEDGEMENTS

This work was supported by a grant from the National Biomedical Research Foundation to Bentley University. The authors thank Prateet Shah for his contributions to this manuscript as well as Drs. Michael Boss and Nancy Hsiung for their constructive suggestions.

## SUPPLEMENTAL FIGURES

**Supplemental Figure 1 A–G. Annual publications for seven vaccine technologies used in candidate COVID-19 vaccines exhibiting logistic or exponential advance fit to the TIME MODEL**. The curve shown represents the first derivative of the exponentiated logistic function. The *established* point represents the point of maximum deceleration of publication activity.

**Supplemental Figure 2 A–G. Cumulative publications for seven vaccine technologies used in candidate COVID-19 vaccines exhibiting logistic or exponential advance fit to the TIME MODEL**. The curve shown represents exponentiated logistic function. Data is shown on a linear scale.

**Supplemental Figure 3 A–G. Cumulative publications for seven vaccine technologies used in candidate COVID-19 vaccines exhibiting logistic or exponential advance fit to the TIME MODEL shown on log scale**. The curve shown represents exponentiated logistic function. Data is shown on a log scale, which illustrates the classic “S-curve” pattern of technological advance seen in other fields of technology.

## REFERENCES

Arregui, Sergio, Sanz, Joaquín, Marinova, Dessislava, Martín, Carlos, and Moreno, Yamir. 2016. On the impact of masking and blocking hypotheses for measuring the efficacy of new tuberculosis vaccines.” PeerJ 4:e1513. doi: 10.7717/peerj.1513

Ball, P. 2020. “Anti-vaccine movement could undermine efforts to end coronavirus pandemic, researchers warn” Nature. doi: 10.1038/d41586-020-01423-4.

Basu, Anirban. 2020. “Estimating the infection fatality rate among symptomatic Covid-19 cases in the United States: Study estimates the COVID-19 infection fatality rate at the US county level” Health Affairs https://doi.org/10.1377/hlthaff.2020.00455

Batson, Amie and Bekier, Matthias M. 2001. “Vaccines where they’re needed” The McKinsey Quarterly. Autumn 2001..

Beierlein, Jennifer M, McNamee, Laura M., and Ledley, Fred D. 2017. “As technologies for nucleotide therapeutics mature, products emerge” Molecular Therapy-Nucleic Acids 9:379–386.

Beierlein, Jennifer M, McNamee, Laura M., Walsh, Michael J., Kaitin, Kenneth I., DiMasi, Joseph A., and Ledley, Fred D. 2017 “Landscape of innovation for cardiovascular pharmaceuticals: From basic science to new molecular entities” Clinical Therapeutics 39(7):1409–1425.

Blume, Stuart S. 2005. “Lock in, the state and vaccine development: Lessons from the history of the polio vaccines” Research Policy 34 (2):159–173.

Boyack, Kevin W. and Jordan, Paul. 2011. “Metrics associated with NIH funding: A high-level view” Journal of the American Medical Informatics Association 18 (4):423–431.

Christensen, Clayton M. 1992. “Exploring the limits of the technology S curve. Part I: component technologies” Production and Operations Management 1 (4):334–357.

Christensen, Clayton M. 1997. The Innovator’s Dilemma: When New Technologies Cause Great Firms to Fail. Boston, MA: Harvard Business School Press.

Clausing, Don and Holmes, Maurice. 2010. “Technology readiness” Research-Technology Management 53 (4):52–59.

Cleary, Ekaterina Galkina, Beierlein, Jennifer M., Khanuja, Navleen Surjit, McNamee, Laura M., and Ledley, Fred D. 2018. “Contribution of NIH funding to new drug approvals 2010–2016” Proceedings of the National Academy of Sciences 115 (10):2329–2334.

Cleary, Ekaterina Galkina, Jackson, Matthew J., Folchman-Wagner, Zoe, and Ledley, Fred D. 2020. Foundational research and NIH funding enabling Emergency Use Authorization of remdesivir for COVID-19.” medRxiv. https://www.medrxiv.org/content/10.1101/2020.07.01.20144576v1

Cleary, Ekaterina Galkina, Jackson, Matthew J., and Ledley, Fred D. 2020. “Government as the First Investor in Biopharmaceutical Innovation: Evidence From New Drug Approvals 2010–2019” INET Economics Working Paper Series, September 2020. https://www.ineteconomics.org/research/research-papers/government-as-the-first-investor-inbiopharmaceutical-innovation-evidence-from-new-drug-approvals-2010-2019

Corey, Lawrence, Mascola, John R., Fauci, Anthony S., and Collins, Francis S. 2020. “A strategic approach to COVID-19 vaccine R&D” Science 368 (6494):948–950.

De Beuckelaer, Ans, Pollard, Charlotte, Van Lint, Sandra, Roose, Kenny, Van Hoecke, Lien, Naessens, Thomas, Udhayakumar, Vimal Kumar, Smet, Muriel, Sanders, Niek, and Lienenklaus, Stefan. 2016. “Type I interferons interfere with the capacity of mRNA lipoplex vaccines to elicit cytolytic T cell responses” Molecular Therapy 24 (11):2012–2020.

Decaro, Nicola, Campolo, Marco, Mari, Viviana, Desario, Costantina, Colaianni, Maria Loredana, Di Trani, Livia, Cordioli, Paolo, and Buonavoglia, Canio. 2009. “A candidate modified-live bovine coronavirus vaccine: Safety and immunogenicity evaluation” The New Microbiologica 32 (1):109–113.

Demertzis, Zachary D., Dagher, Carina, Malette, Kelly M. Fadel, Raef A., Bradley, Patrick B., Brar, Indira, Rabbani, Bobak T., and Suleyman, Geehan. 2020. “Cardiac Sequelae of Novel Coronavirus Disease 2019 (COVID-19): A Clinical Case Series” European Heart Journal-Case Reports doi.org/10.1093/ehjcr/ytaa179.

Fiani, Brian, Covarrubias, Claudia, Desai, Aditi, Sekhon, Manraj, and Jarrah, Rayan. 2020. “A contemporary review of neurological sequelae of COVID-19” Frontiers in Neurology 11:640.

Flaxman, Seth, Mishra, Swapnil, Gandy, Axel, Unwin, H., Coupland, Helen, Mellan, T., Zhu, Harisson, Berah, Tresnia, Eaton, J., and Guzman, P.Perez. 2020. “Report 13: Estimating the number of infections and the impact of non-pharmaceutical interventions on COVID-19 in 11 European countries” https://www.imperial.ac.uk/mrc-global-infectious-diseaseanalysis/covid-19/report-13-europe-npi-impact/.

Folegatti, Pedro M., Bittaye, Mustapha, Flaxman, Amy, Lopez, Fernando Ramos, Bellamy, Duncan, Kupke, Alexandra, Mair, Catherine, Makinson, Rebecca, Sheridan, Jonathan, and Rohde, Cornelius. 2020. “Safety and immunogenicity of a candidate Middle East respiratory Ssyndrome coronavirus viral-vectored vaccine: A dose-escalation, open-label, non-randomised, uncontrolled, Phase 1 trial” The Lancet Infectious Diseases doi.org/10.1016/S1473-3099(20)30160-2,

Foster, Richard N. 1982. “Effective R&D operations in the’80s: boosting the payoff from R&D” Research Management 25 (1):22–27.

Francis, Donald P. 2010. “Successes and failures: Worldwide vaccine development and application” Biologicals 38 (5):523–528.

GAO. 1999. “BEST PRACTICES: Better Management of Technology Development Can Improve Weapon System Outcomes” General Accounting Office. https://www.gao.gov/products/NSIAD-99-162

Gouglas, Dimitrios, Le, Tung Thanh, Henderson, Klara, Kaloudis, Aristidis, Danielsen, Trygve, Hammersland, Nicholas Caspersen, Robinson, James M., Heaton, Penny M., and Røttingen, John-Arne. 2018. “Estimating the cost of vaccine development against epidemic infectious diseases: A cost minimisation study” The Lancet Global Health 6(12):e1386–e1396.

Harapan, Harapan, Michie, Alice, Sasmono, R.Tajo, and Imrie, Allison. 2020. “Dengue: A minireview.” Viruses 12(8):829.

KFF.2020. The U.S. and Gavi, the Vaccine Alliance. Kaiser Family Foundation. https://www.kff.org/global-health-policy/fact-sheet/the-u-s-and-gavi-the-vaccine-alliance/, June 2020.

Kim, Hyunsuh, Webster, Robert G., and Webby, Richard J. 2018. “Influenza virus: Dealing with a drifting and shifting pathogen” Viral Immunology 31 (2):174–183.

Kristensen, Debra D, Lorenson, Tina, Bartholomew, Kate, and Villadiego, Shirley.2016 “Can thermostable vaccines help address cold-chain challenges? Results from stakeholder interviews in six low-and middle-income countries” Vaccine 34 (7):899–904.

Lambert, Paul-Henri, Ambrosino, Donna M., Andersen, Svein R., Baric, Ralph S., Black, Steven B., Chen, Robert T., Dekker, Cornelia L., Didierlaurent, Arnaud M., Graham, Barney S., and Martin, Samantha D. 2020. “Consensus summary report for CEPI/BC March 12–13, 2020 meeting: Assessment of risk of disease enhancement with COVID-19 vaccines” Vaccine. doi: 10.1016/j.vaccine.2020.05.064 May 2020.

Laurens, Matthew B. 2018. “The promise of a malaria vaccine—Are we closer?” Annual Review of Microbiology 72:273–292.

Ledley, F.D., McNamee, L.M., Uzdil, V., and Morgan, I.W. 2014. “Why commercialization of gene therapy stalled; Examining the life cycles of gene therapy technologies” Gene Therapy 21 (2):188–194.

Li, Jianping, Helal, Zeinab H., Karch, Christopher P., Mishra, Neha, Girshick, Theodore, Garmendia, Antonio, Burkhard, Peter, and Khan, Mazhar I. 2018. “A self-adjuvanted nanoparticle based vaccine against infectious bronchitis virus” PloS ONE 13(9):e0203771.

McNamee, Laura M. and Ledley, Fred D. 2012. “Patterns of technological innovation in biotech” Nature Biotechnology 30 (10):937–943.

McNamee, Laura M. and Ledley, Fred D. 2017. “Modeling timelines for translational science in cancer; The impact of technological maturation” PLoS ONE 12 (3):e0174538.

McNamee, Laura M, Walsh, Michael Jay, and Ledley, Fred D. 2017 “Timelines of translational science: From technology initiation to FDA approval” PLoS ONE 12 (5):e0177371.

McPherson, Clifton, Chubet, Richard, Holtz, Kathy, Honda-Okubo, Yoshikazu, Barnard, Dale, Cox, Manon, and Petrovsky, Nikolai. 2016. “Development of a SARS coronavirus vaccine from recombinant spike protein plus delta inulin adjuvant” Methods Mol Biol 1403:269–84.

Mehand, Massinissa Si, Al-Shorbaji, Farah, Millett, Piers, and Murgue, Bernadette. 2018. “The WHO R&D blueprint: 2018 review of emerging infectious diseases requiring urgent research and development efforts” Antiviral Research 159:63–67.

Modjarrad, Kayvon. 2016. “MERS-CoV vaccine candidates in development: The current landscape” Vaccine 34 (26):2982–2987.

Modjarrad, Kayvon, Roberts, Christine C., Mills, Kristin T., Castellano, Amy R., Paolino, Kristopher, Muthumani, Kar, Reuschel, Emma L., Robb, Merlin L., Racine, Trina, and Oh, Myoung-don. 2019. “Safety and immunogenicity of an anti-Middle East respiratory syndrome coronavirus DNA vaccine: a Phase 1, open-label, single-arm, dose-escalation trial” The Lancet Infectious Diseases 19 (9):1013–1022.

NSF. 2018. Definitions of Research and Development: An Annotated Compilation of Official Sources. https://www.nsf.gov/statistics/randdef/rd-definitions.pdf, March 2018.

NIH. 2019. Estimates of Funding for Various Research, Condition, and Disease Categories (RCDC). https://exporter.nih.gov/ExPORTER_Catalog.aspx

NIH. 2020. Research portfolio online reporting tools (RePORTER), ExPORTER data catalog.

NVAC (National Vaccine Advisory Committee). 1997. “United States vaccine research: a delicate fabric of public and private collaboration.” Pediatrics 100 (6):1015–1020.

O’Callaghan, Kevin P, Blatz, Allison M., and Offit, Paul A. 2020. “Developing a SARS-CoV-2 vaccine at warp speed.” JAMA 2020;324(5):437–438.

Pardi, Norbert, Hogan, Michael J., Porter, Frederick W., and Weissman, Drew. 2018. “mRNA vaccines—a new era in vaccinology.” Nature reviews Drug Discovery 17 (4): 261–279.

Plotkin, Stanley. 2014. “History of vaccination.” Proceedings of the National Academy of Sciences 111 (34):12283–12287.

>Plotkin, Stanley A. 2017. “Vaccines for epidemic infections and the role of CEPI” Human Vaccines & Immunotherapeutics 13 (12):2755–2762.

Porter, Kevin R. and Raviprakash, Kanakatte. 2017. “DNA vaccine delivery and improved immunogenicity.” Current Issues in Molecular Biology 22:129–138.

Pronker, Esther S., Weenen, Tamar C., Commandeur, Harry, Claassen Eric, H.J.H.M., and Osterhaus, Albertus D.M.E. 2013. “Risk in vaccine research and development quantified.” PloS ONE 8(3):e57755. doi.org/10.1371/journal.pone.0057755.

Randolph, Haley E. and Barreiro, Luis B. 2020. “Herd immunity: Understanding COVID-19.” Immunity 52 (5):737–741.

Rappuoli, Rino, Black, Steven, and Bloom, David E. 2019. “Vaccines and global health: In search of a sustainable model for vaccine development and delivery.” Science Translational Medicine 11 (497):eaaw2888. doi: 10.1126/scitranslmed.aaw2888.

Safrit, Jeffrey T., Fast, Patricia E., Gieber, Lisa, Kuipers, Hester, Dean Hansi J., and Koff, Wayne C. 2016. “Status of vaccine research and development of vaccines for HIV-1” Vaccine 34 (26):2921–2925.

Schlake, Thomas, Thess, Andreas, Fotin-Mleczek, Mariola, and Kallen, Karl-Josef. 2012. “Developing mRNA-vaccine technologies.” RNA Biology 9 (11):1319–1330.

Stephens, Peter. 2014. “Vaccine R&D: past performance is no guide to the future” Vaccine 32(19):2139–2142.

Tau, Noam, Yahav, Dafna, and Shepshelovich, Daniel. 2020. “Postmarketing safety of vaccines approved by the US Food and Drug Administration: A cohort study” Annals of Internal Medicine. doi.org/10.7326/M20-2726

Thomas, Stephen J. and Barrett, Alan. 2020. “Zika vaccine pre-clinical and clinical data review with perspectives on the future development” Human Vaccines & Immunotherapeutics:1–13. doi: 10.1080/21645515.2020.1730657.

WHO. 2016. “An R&D blueprint for action to prevent epidemics” World Health Organization, https://www.who.int/blueprint/what/improving-coordination/workstream_5_document_on_financing.pdf?ua=1

WHO. 2018. “Dengue vaccine: WHO position paper, September 2018—recommendations” Vaccine 37:4848–4849.

WHO. 2019. “Global vaccine market report” World Health Organization. https://apps.who.int/iris/handle/10665/311278

WHO. 2020. “DRAFT landscape of COVID-19 candidate vaccines” World Health Organization. https://www.who.int/publications/m/item/draft-landscape-of-covid-19-candidatevaccines (Accessed July 31, 2020)

Zhu, Na, Zhang, Dingyu, Wang, Wenling, Li, Xingwang, Yang, Bo, Song, Jingdong, Zhao, Xiang, Huang, Baoying, Shi, Weifeng, and Lu, Roujian. 2020. “A novel coronavirus from patients with pneumonia in China, 2019” New England Journal of Medicine. 382:727–733

